# Heterogeneity in Preclinical Alzheimer’s Disease Trial Cohort Identified by Image-based Data-Driven Disease Progression Modelling

**DOI:** 10.1101/2023.02.07.23285572

**Authors:** Cameron Shand, Pawel J. Markiewicz, David M. Cash, Daniel C. Alexander, Michael C. Donohue, Frederik Barkhof, Neil P. Oxtoby, the Alzheimer’s Disease Neuroimaging Initiative

**Affiliations:** Centre for Medical Image Computing, Department of Computer Science, University College London, London, UK; Centre for Medical Image Computing, Department of Medical Physics and Biomedical Engineering, University College London, London, UK; Dementia Research Centre, UCL Queen Square Institute of Neurology, University College London, London, UK; Alzheimer’s Therapeutic Research Institute, Keck School of Medicine, University of Southern California, San Diego, USA; Department of Radiology and Nuclear Medicine, VU University Medical Center, Amsterdam, Netherlands

**Author notes:** Corresponding author: Neil Oxtoby.

## Abstract

**Importance:** Undetected biological heterogeneity adversely impacts trials in Alzheimer’s disease because rate of cognitive decline — and perhaps response to treatment — differs in subgroups. Recent results show that data-driven approaches can unravel the heterogeneity of Alzheimer’s disease progression. The resulting stratification is yet to be leveraged in clinical trials.

**Objective:** Investigate whether image-based data-driven disease progression modelling could identify baseline biological heterogeneity in a clinical trial, and whether these subgroups have prognostic or predictive value.

**Design:** Screening data from the Anti-Amyloid Treatment in Asymptomatic Alzheimer Disease (A4) Study collected between April 2014 and December 2017, and longitudinal data from the Alzheimer’s Disease Neuroimaging Initiative (ADNI) observational study downloaded in February 2022 were used.

**Setting:** The A4 Study is an interventional trial involving 67 sites in the US, Canada, Australia, and Japan. ADNI is a multi-center observational study in North America.

**Participants:** Cognitively unimpaired amyloid-positive participants with a 3-Tesla T1-weighted MRI scan. Amyloid positivity was determined using florbetapir PET imaging (in A4) and CSF Aβ(1-42) (in ADNI).

**Main Outcomes and Measures:** Regional volumes estimated from MRI scans were used as input to the Subtype and Stage Inference (SuStaIn) algorithm. Outcomes included cognitive test scores and SUVr values from florbetapir and flortaucipir PET.

**Results:** We included 1,240 Aβ+ participants (and 407 Aβ− controls) from the A4 Study, and 731 A4-eligible ADNI participants. SuStaIn identified three neurodegeneration subtypes — *Typical, Cortical, Subcortical* — comprising 523 (42%) individuals. The remainder are designated subtype zero (insufficient atrophy). Baseline PACC scores (A4 primary outcome) were significantly worse in the *Cortical* subtype (median = -1.27, IQR=[-3.34,0.83]) relative to both subtype zero (median=-0.013, IQR=[-1.85,1.67], P<.0001) and the *Subcortical* subtype (median=0.03, IQR=[-1.78,1.61], P=.0006). In ADNI, over a four-year period (comparable to A4), greater cognitive decline in the mPACC was observed in both the *Typical* (−0.23/yr; 95% CI, [-0.41,-0.05]; P=.01) and *Cortical* (−0.24/yr; [-0.42,-0.06]; P=.009) subtypes, as well as the CDR-SB (*Typical*: +0.09/yr, [0.06,0.12], P<.0001; and *Cortical*: +0.07/yr, [0.04,0.10], P<.0001).

**Conclusions and Relevance:** In a large secondary prevention trial, our image-based model detected neurodegenerative heterogeneity predictive of cognitive heterogeneity. We argue that such a model is a valuable tool to be considered in future trial design to control for previously undetected variance.

**Key Points:** *Question:* Can MRI-based computational subtypes of preclinical neurodegeneration predict cognitive outcomes?

*Findings:* In this cross-sectional analysis of magnetic resonance imaging (MRI) data at screening (pre-randomization) in the preclinical Anti-Amyloid Treatment in Asymptomatic Alzheimer disease (A4) Study, we detected considerable neurodegenerative heterogeneity using data-driven disease progression modelling. The MRI-based computational subtypes identified by Subtype and Stage Inference (SuStaIn) differed in baseline cognitive test scores (A4) and in longitudinal cognitive decline (ADNI), with sufficient heterogeneity to potentially obscure treatment effect in A4 trial outcomes.

*Meaning:* Data-driven disease progression modelling of screening MRI scans can predict heterogeneity in cognitive performance/decline and potentially reduce heterogeneity in future clinical trials.

## Introduction

There is increasing evidence of biological heterogeneity to accompany the observed clinical heterogeneity in Alzheimer’s disease (AD), highlighting a range of potential disease subtypes and disease trajectories^1–6^. This heterogeneity reflects the inherent complexity of the disease and may have contributed to the high failure rate of clinical trials^7^, where study cohorts likely contain individuals potentially belonging not only to different subtypes, but also being at different stages of biological severity (even within a clinical/biomarker-based group), resulting in heterogeneous rates of progression independent of treatment effect^8,9^. This compounds the already difficult task of identifying treatment effects in AD clinical trials, where neuropsychological test scores or single biomarkers (e.g. amyloid PET SUVr) are used as outcomes^10,11^. Nearly half of currently ongoing Phase 3 AD trials (for disease-modifying therapies) did not use biomarkers as inclusion criteria, relying instead on neuropsychological tests^10^. Although some composite neuropsychological tests are sensitive to relatively early, subtle changes in cognitive function^12^, underlying pathology in the preclinical phase cannot be fully characterized without biomarkers^12,13^.

Recognizing this limitation in trial designs, biomarkers are increasingly used in screening protocols^10,11,14,15^, yet still incur high failure rates likely due to the limited temporal resolution of a single biomarker, and the presence of undetected subtypes having differing cognitive trajectories/prognoses^4,16^. Data-driven methods such as disease progression modelling (DPM) have previously characterized heterogeneity (both in variation of severity and the presence of subtypes) typically using image-based input features^5,17^. The ability to utilize high-dimensional medical imaging data enables deeper comparisons between individuals beyond univariate cut-offs, providing utility in assessing heterogeneity across a cohort. In this study, we use DPM to identify undetected MRI-based heterogeneity at screening in the Anti-Amyloid Treatment in Asymptomatic AD (A4) Study^18^, analyze these subtypes for differences in trial outcome measures (at baseline), and investigate differences across subtypes in longitudinal outcomes in the Alzheimer’s Disease Neuroimaging Initiative (ADNI)^19^ observational study to characterize the potential impact such heterogeneity may have on the A4 interventional trial outcomes.

## Methods

Our primary aim is to test whether biological heterogeneity is predictive of cognitive decline in the carefully selected A4 Study clinical trial cohort. We train a subtyping DPM on imaging (MRI) data, then compare cognitive outcomes across subtypes cross-sectionally (in A4) and longitudinally (in ADNI).

### Participants

Data comes from cognitively unimpaired participants in the A4 Study interventional trial (NCT02008357) and the ADNI observational study (eMethods 1 in supplementary material). The A4 Study is a prevention trial investigating whether solanezumab reduces cognitive decline in an Aβ+ preclinical cohort^14,20^, with the primary outcome being the change from baseline of the Preclinical Alzheimer Cognitive Composite (PACC)^21^. Secondary outcomes include changes from baseline in the Clinical Dementia Rating Sum of Boxes (CDR-SB) score and the Cognitive Function Index (CFI) total score (combining participant and study partner scores)^22^. ADNI is an observational study investigating biomarker and clinical changes in individuals diagnosed with, or at-risk of, AD.

The A4 Study selected individuals using the following criteria^14^: Mini-Mental State Examination (MMSE) score of 25-30; Clinical Dementia Rating (CDR) global score of 0; Logical Memory Delayed Recall (LMDR-IIa) score of 6-18; and, amyloid positivity (Aβ+). Amyloid positivity was defined following florbetapir PET imaging, using a mean cortical standardized uptake value ratio (SUVr) with a whole cerebellar reference region of ≥1.15 (or ≥1.10 on a positive visual read with a 2-reader consensus). To maximize data availability in ADNI, CSF Aβ(1-42) was instead used to determine amyloid positivity using a cut-off of 977pg/mL^23^. Cognitively unimpaired individuals in ADNI were selected using the same inclusion criteria as the A4 Study. ADNI data was downloaded from the LONI Image & Data Archive on 27th February 2022.

Control subjects for our study were Aβ− and APOE ε4 non-carriers from the A4 Study. These controls (N=407) were used for covariate adjustment and as a normative reference when analyzing both A4 and ADNI cohorts (see below).

### Data Pre-Processing

All available 3-Tesla (3T) T1-weighted MRI were obtained for both the A4 and ADNI cohorts. Images from ADNI were selected having passed overall quality control. All images were processed using FreeSurfer v7.1.1^24^ within a publicly available containerized pipeline (https://github.com/e-dads/freesurfer), with cortical and subcortical volumes combined into bilateral averages in 13 regions of interest^25^. Original image acquisition protocols for both studies can be found in their respective documentation.

Prior to input into our DPM, FreeSurfer volumes were adjusted for healthy/normal linear trends (in controls) in age, years of education, sex, and intracranial volume, estimated using a general linear model^26^. The covariate-adjusted volumes were transformed into z-scores relative to controls.

The amyloid (florbetapir) and tau (flortaucipir) PET images acquired in the A4 Study were analyzed to investigate differences across subtypes. For this analysis, we used a newly-developed multi-platform software AmyPET (https://github.com/AMYPAD/AmyPET), extending NiftyPET^27^. AmyPET enables robust quantification of PET scans while using SPM12 for core MR-PET image registration^28^, estimating amyloid/tau load with high quantitative accuracy and precision from raw count PET data to accurately account for common head motion. Protocol details for amyloid and tau PET acquisition are provided in eMethods 2 (supplementary material).

### Disease Progression Modelling

To characterize biological heterogeneity in the A4 Study cohort, we used the Subtype and Stage Inference (SuStaIn) algorithm^5^ as our DPM. This approach combines clustering with event-based modelling^17,29^ to reconstruct one (or more) sequences of disease progression from cross-sectional data, simultaneously disentangling pathological and temporal heterogeneity to identify both disease subtype(s) and severity (i.e. stage) across a cohort. SuStaIn has been applied to identify image-based subtypes and spreading patterns in AD^5,16,30^, other forms of dementia such as frontotemporal dementia^31^, and other neurodegenerative diseases such as multiple sclerosis^32^.

Disease “events” (transitions of a regional volume to abnormality) are defined by reaching progressive z-scores of 1 and 2 (relative to controls), from which SuStaIn determines the event sequence that maximizes the data likelihood. SuStaIn then iteratively determines multiple such sequences, referred to here as disease subtypes, that maximize the likelihood for subsets of the data from one subtype up to a pre-defined maximum (set as *N*_*max*_ = 4), with the optimal number of subtypes selected using cross-validation (described below). Markov chain Monte Carlo (MCMC) sampling is used to estimate uncertainty in the sequence(s).

SuStaIn was trained using the 1,240 Aβ+ participants with complete MRI data from the A4 Study. This produced a model for each number of subtypes investigated. To select a single model, we performed 10-fold cross-validation and calculated the cross-validation information criterion (CVIC)^13^, which balances minimizing model complexity (i.e. fewer subtypes) and maximizing how well the model fits the data. The model with the minimal CVIC is selected, and then used to assign both a maximum-likelihood subtype and stage to each participant (for both A4 and ADNI cohorts)^5,17^.

### Statistical Analyses

The original A4 Study analysis^14^ found significant differences between the Aβ+ and Aβ− groups at baseline for both the PACC and CFI scores, with the Aβ+ group performing worse on both scores. We performed a similar analysis stratified by DPM subtype assignment, using Pearson’s χ^2^ (adjusted for multiple comparisons using Holm-Bonferroni correction). This analysis was repeated for the ADNI subset to compare differences between the subtypes, and between the A4 and ADNI cohorts to identify any systematic differences in demographic and cognitive variables common to both studies.

Associations between DPM subtypes and cognitive outcomes are assessed for (pairwise) differences using a two-tailed Mann-Whitney U test (with Holm-Bonferroni correction). This was performed cross-sectionally in A4 using the PACC and CFI (total) scores to potentially identify undetected baseline cognitive heterogeneity revealed by the DPM.

We then analyzed longitudinal cognitive trajectories in ADNI for two similar trial outcomes available in ADNI: a modified version of the PACC (mPACC), which uses the (log-transformed) Trail-Making Test B to replace the unavailable Digit Symbol Substitution test^33^, and the CDR-SB. The impact of subtype assignment and resulting differences in cognitive decline was analyzed using a linear mixed effects model, including age at baseline, time since baseline (continuous), and subtype-by-time interactions as fixed effects, as well as allowing for participant-specific random intercepts^34^. To mirror the A4 Study, we restricted this analysis using data obtained over a similar period of observation (4 years from baseline).

## Results

### A4 Study MRI-based Disease Progression Subtypes

A total of 1,240 Aβ+ participants with complete MRI data were used as input into SuStaIn, following covariate adjustment and z-scoring using the 407 Aβ− screen-failures. Cross-validation indicated that the 3-subtype model best fit the data. Table 1 shows demographic and cognitive score variables for the Aβ− control group (for reference), the whole Aβ+ group, and the Aβ+ subtype groups which we designate as *Typical, Cortical*, and *Subcortical* based on the observed patterns of atrophy (Figure 1). There were no significant differences in demographic variables between the subtypes nor in the CDR-SB or MMSE, but the PACC, CFI (total), and Digit Symbol Substitution test scores were significantly different across the subtypes.

**Table 1.** Characteristics of the A4 Study cohort, separated between Aβ− (control) and Aβ+ (SuStaIn input) groups, further separated by subtype assignment. There were no significant differences among subtypes in demographic variables, the CDR-SB and the MMSE according to either an ANOVA or Pearson’s χ^2^ test (following Holm-Bonferroni adjustment for multiple comparisons). The PACC, CFI (total), and Digital Symbol Substitution test scores showed significant (P < .001) differences between subtypes. Abbreviations: SUVr – Standardized Uptake Value ratio; PACC – Preclinical Alzheimer Cognitive Composite; CFI – Cognitive Function Index; MMSE – Mini-Mental State Examination; CDR-SB – Clinical Dementia Rating Sum of Boxes

| Characteristic | Aβ- (Control group) | Aβ+ (Whole group) | Subtype Zero (sub-threshold) | Typical Subtype | Cortical Subtype | Subcortical Subtype | Adjusted P-value (across subtypes) |
| --- | --- | --- | --- | --- | --- | --- | --- |
| No. individuals | 407 | 1240 | 717 | 170 | 179 | 174 |  |
| Age, yrs, mean (SD) | 70.8 (4.4) | 72.0 (4.9) | 71.9 (4.6) | 72.4 (5.3) | 72.6 (5.2) | 71.6 (4.9) | .58<sup>a</sup> |
| Female, (%) | 258 (63.4) | 730 (59.0) | 445 (62.1) | 98 (57.6) | 103 (57.5) | 84 (48.3) | .08<sup>b</sup> |
| Education, yrs, mean (SD) | 16.7 (2.5) | 16.6 (2.8) | 16.6 (2.8) | 16.0 (2.8) | 16.6 (2.8) | 16.8 (2.9) | .18<sup>a</sup> |
| Family history of dementia (%) | 249 (61.2) | 924 (74.5) | 525 (73.2) | 130 (76.5) | 136 (76.0) | 133 (76.4) | .58<sup>b</sup> |
| APOE ε4 alleles (%) | | | | | | | |
| 0 | 407 (100.0) | 506 (40.8) | 297 (41.4) | 62 (36.5) | 73 (40.8) | 74 (42.5) | .68<sup>b</sup> |
| 1 | 0 (0.0) | 621 (50.1) | 364 (50.8) | 92 (54.1) | 82 (45.8) | 83 (47.7) |  |
| 2 | 0 (0.0) | 101 (8.2) | 49 (6.8) | 12 (7.1) | 23 (12.9) | 17 (9.8) |  |
| Missing | 0 (0.0) | 12 (1.0) | 7 (1.0) | 4 (2.4) | 1 (0.6) | 0 (0.00) |  |
| PET SUVR, mean (SD) | 0.99 (0.07) | 1.33 (0.18) | 1.32 (0.17) | 1.35 (0.19) | 1.35 (0.19) | 1.33 (0.18) | .58<sup>a</sup> |
| PACC score, mean (SD) | 0.22 (2.38) | -0.42 (2.69) | -0.15 (2.57) | -0.89 (2.72) | -1.30 (3.04) | -0.13 (2.49) | < .001<sup>a</sup> |
| CDR-SB score, mean (SD) | 0.05 (0.16) | 0.06 (0.17) | 0.05 (0.15) | 0.08 (0.19) | 0.06 (0.16) | 0.07 (0.22) | .50<sup>a</sup> |
| MMSE score, mean (SD) | 28.93 (1.13) | 28.74 (1.28) | 28.80 (1.23) | 28.55 (1.38) | 28.65 (1.41) | 28.78 (1.21) | .50<sup>a</sup> |
| Digit Symbol, mean (SD) | 44.21 (8.85) | 42.59 (8.93) | 43.52 (8.91) | 41.08 (9.19) | 40.55 (8.82) | 42.29 (8.39) | < .001<sup>a</sup> |
| CFI total score, mean (SD) | 2.92 (3.07) | 3.82 (3.49) | 3.43 (3.10) | 4.29 (3.83) | 4.81 (4.17) | 3.93 (3.68) | < .001<sup>a</sup> |

<sup>a</sup> ANOVA
<sup>b</sup> Pearson's $\chi^2$

**Figure 1.**
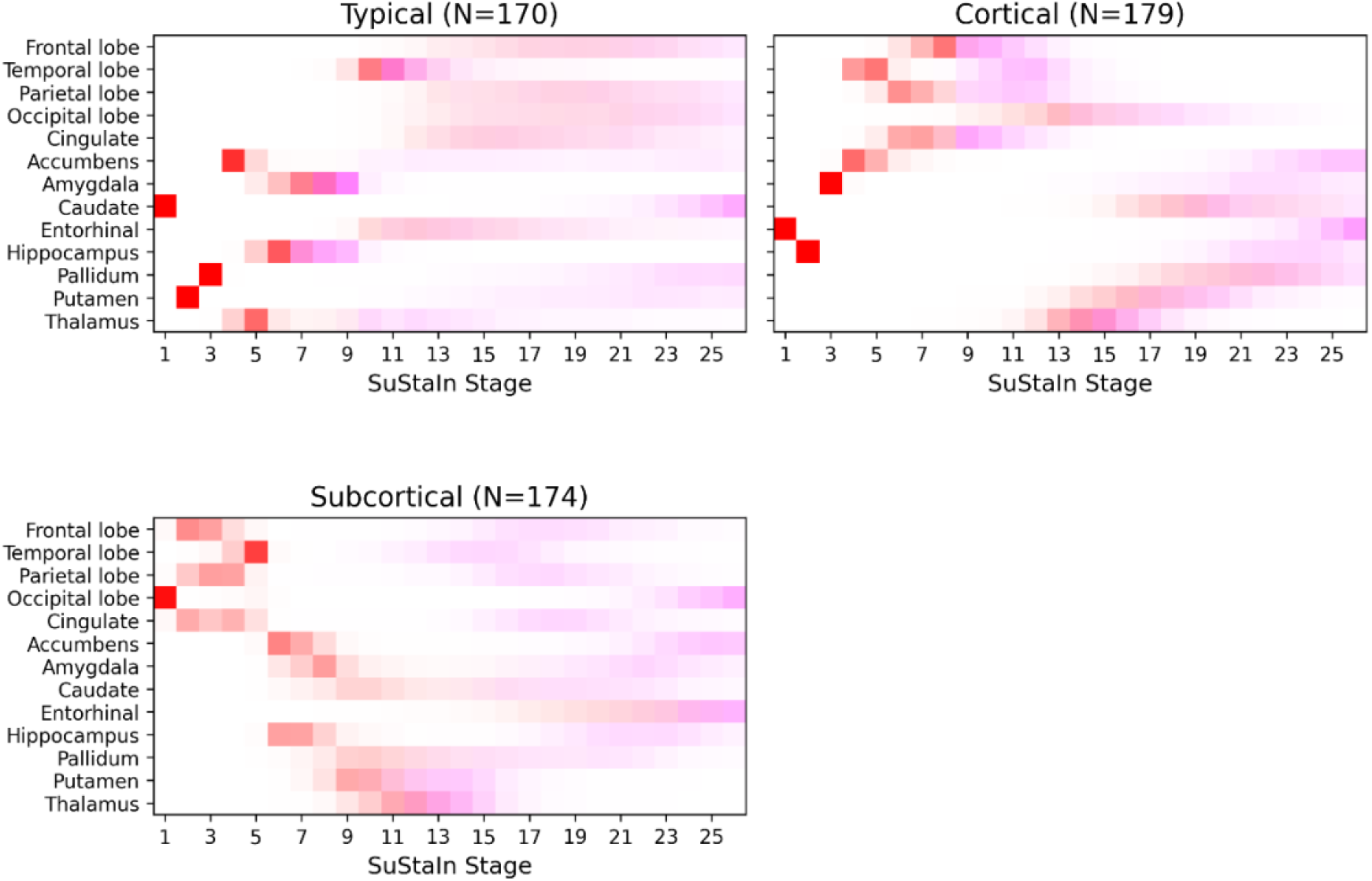
Positional variance diagram showing the disease progression sequences of the final 3-subtype disease progression model trained on the A4 Aβ+ cohort. The red squares indicate a z-score 1 event (abnormality), and magenta squares indicate a z-score 2 event (atrophy). Color intensity is proportional to positional confidence/certainty of the model.

Figure 1 shows a positional variance diagram illustrating the MCMC-sampled posterior distributions for each subtype in the final 3-subtype SuStaIn model. The vertical axis of brain volumes is ordered from cortical to subcortical regions, top-to-bottom. Z-score events along the horizontal axis are color-coded, with z=1 indicating subtle abnormality (in red), and z=2 indicating atrophy (in magenta). Subtypes were named according to the earliest atrophy events (z=2, magenta). In Figure 1, the *Typical* subtype shows early atrophy in the hippocampus, amygdala, and temporal lobe, alongside early yet subtle abnormality in the caudate, putamen, and pallidum. The *Cortical* subtype shows early atrophy in cortical regions (including the cingulate gyrus) and subtle abnormality in the entorhinal cortex, hippocampus, and amygdala. The *Subcortical* subtype shows early atrophy in the putamen and thalamus, albeit with high uncertainty (possibly indicating slow atrophy), alongside subtle abnormality in cortical lobes.

Figure 2 shows the distribution of stage assignments stratified by subtype in the A4 cohort (upper panel) and ADNI cohort (lower panel). Across A4, 523 (42.2%) individuals were assigned to a subtype: 170 (32.5%) *Typical*, 179 (34.2%) *Cortical*, and 174 (33.3%) *Subcortical*. The remaining 717 (57.8%) individuals were not assigned a subtype due to all regional volumes scoring z<1. We refer to these as subtype zero. The *Typical, Cortical*, and *Subcortical* subtypes have median stage assignments of 2 (IQR=1–3), 3 (IQR=1–5), and 3 (IQR=1–5) respectively, consistent with having subtle abnormality reflective of their cognitively unimpaired status.

**Figure 2.**
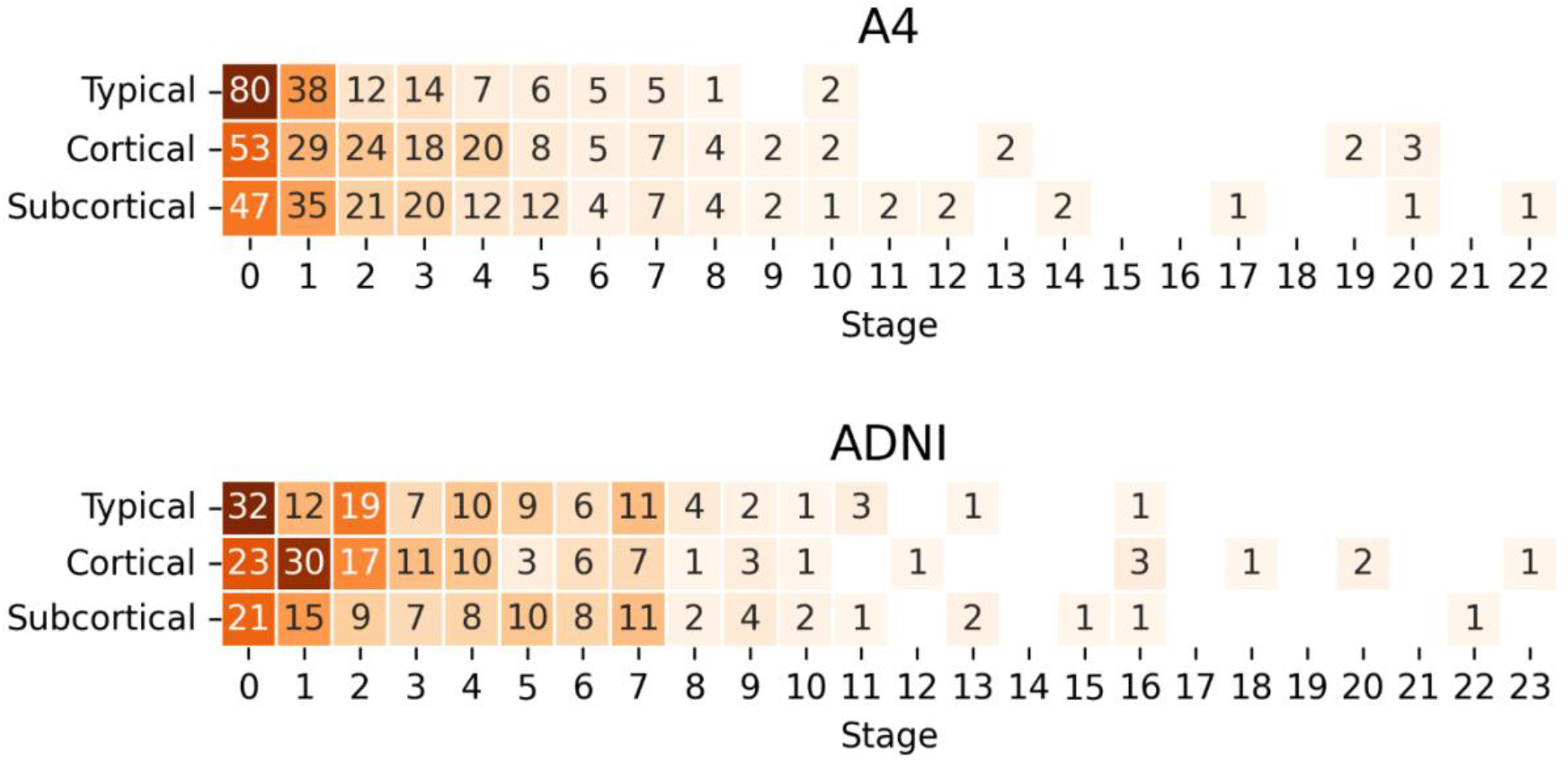
Heatmap and counts of stage assignments per subtype for the A4 Study cohort (upper panel) and ADNI cohort (lower panel).

Mean cortical SUVr used in the A4 Study screening did not show any significant differences across subtypes (Table 1). In Figure 3, our image processing using AmyPET (see Methods) shows SUVr (with a whole cerebellar reference region) values for amyloid (N=1234) and tau (N=392) PET imaging stratified by subtype. Following multiple comparison correction, there was a significant difference between subtype zero and the *Cortical* subtype in amyloid SUVr (P=.04), though the *Cortical* subtype also had a notably higher tau burden (median=1.12; IQR=1.03– 1.16) relative to subtype zero (median=1.08; IQR=1.04–1.12), the *Typical* subtype (median=1.08; IQR=1.05–1.11), and the *Subcortical* subtype (median=1.09; IQR=1.05–1.13).

**Figure 3.**
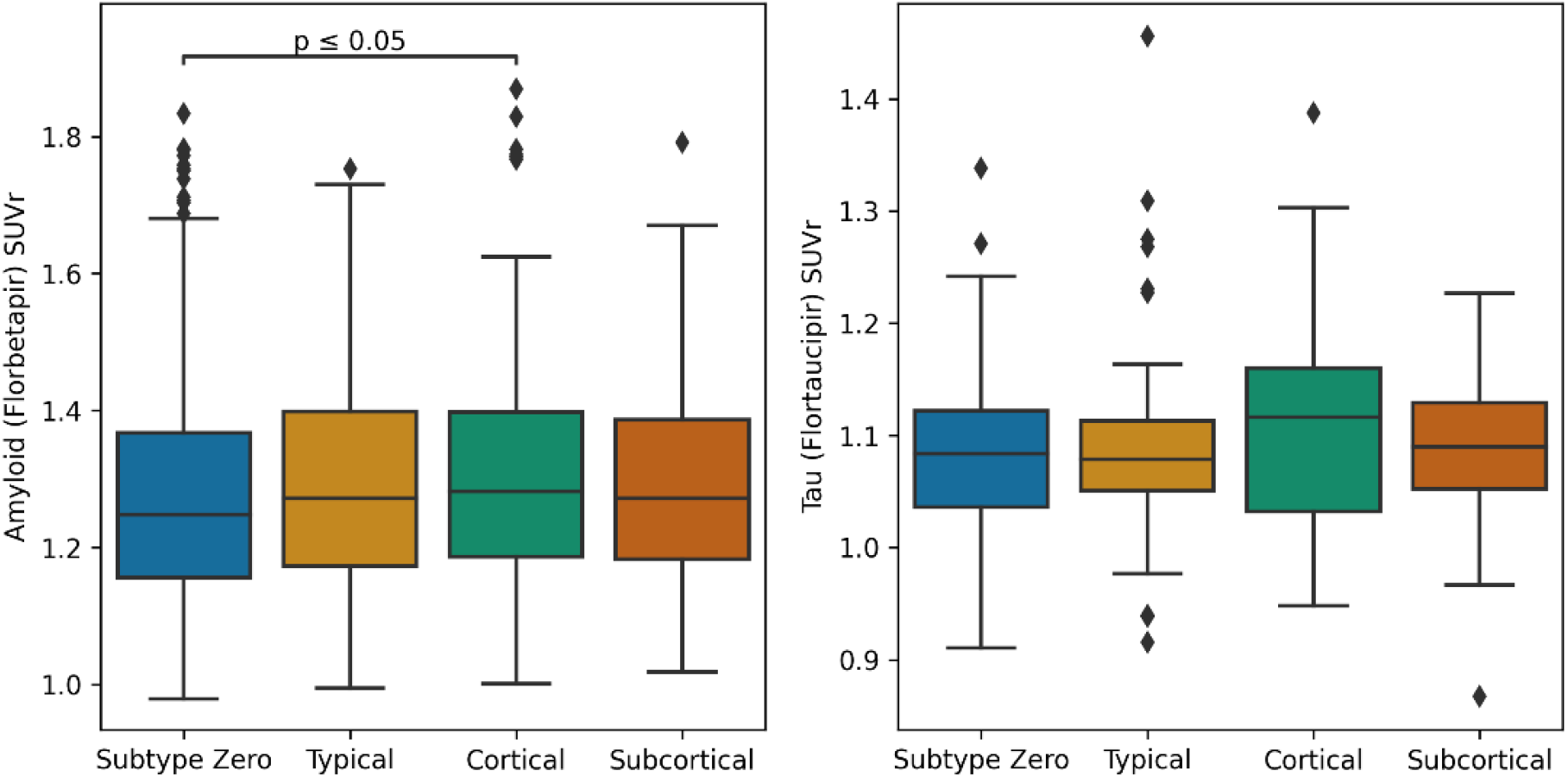
Boxplots showing SUVr values (stratified by model-based subtype) for amyloid (left) and tau (right) PET scans, each using the whole cerebellum as a reference region. Lines are added to show pairwise significant differences (two-tailed Mann-Whitney U test, adjusted for multiple comparisons). Abbreviations: SUVr – standardized uptake value ratio

**Figure 4.**
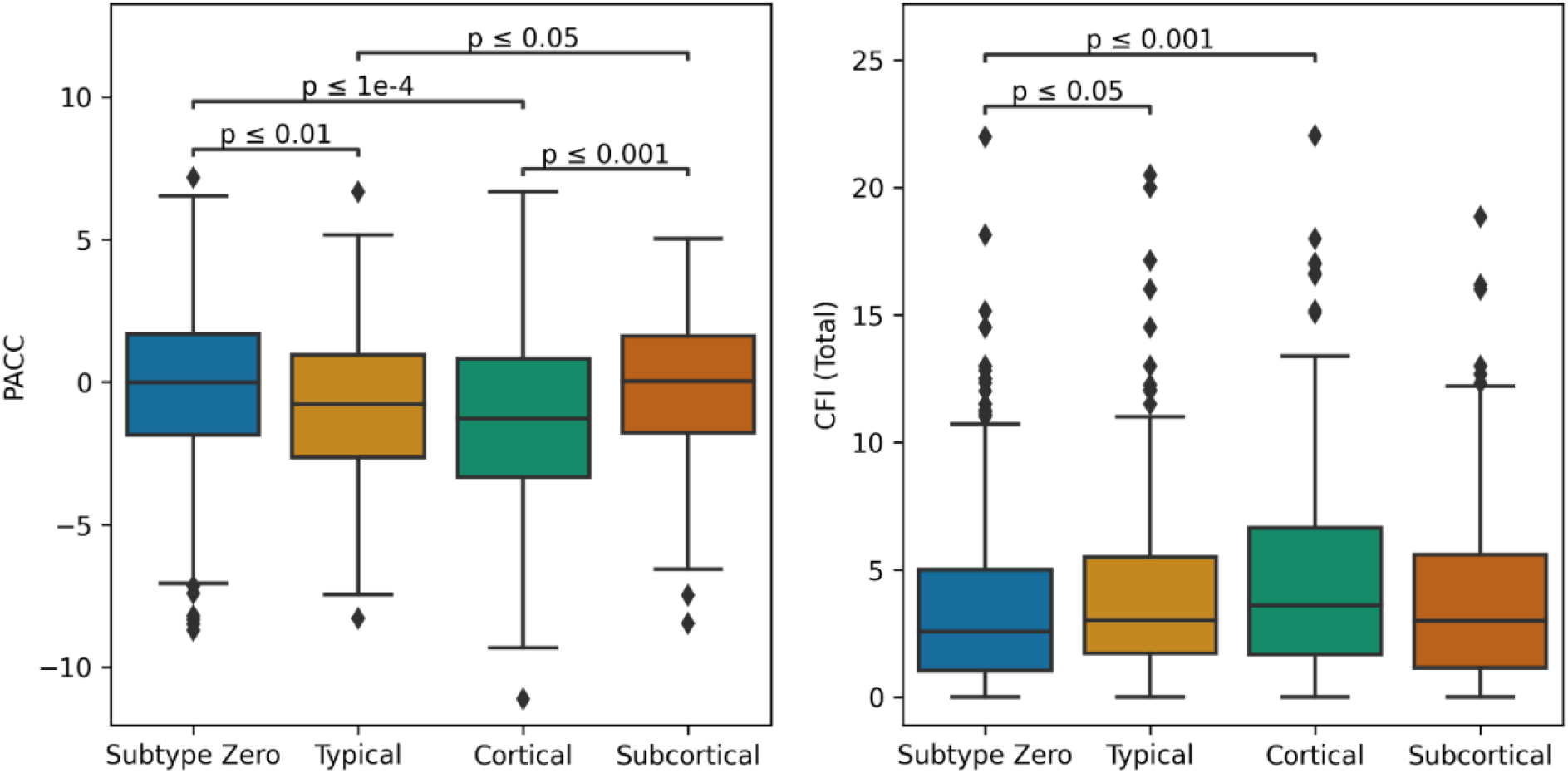
Boxplots of PACC (left) and CFI Total (right) scores in the A4 Study cohort, stratified by model-based subtype. Lines are added to show pairwise significant differences (two-tailed Mann-Whitney U test, adjusted for multiple comparisons). Abbreviations: PACC – Preclinical Alzheimer Cognitive Composite; CFI – Cognitive Function Index.

**Figure 5.**
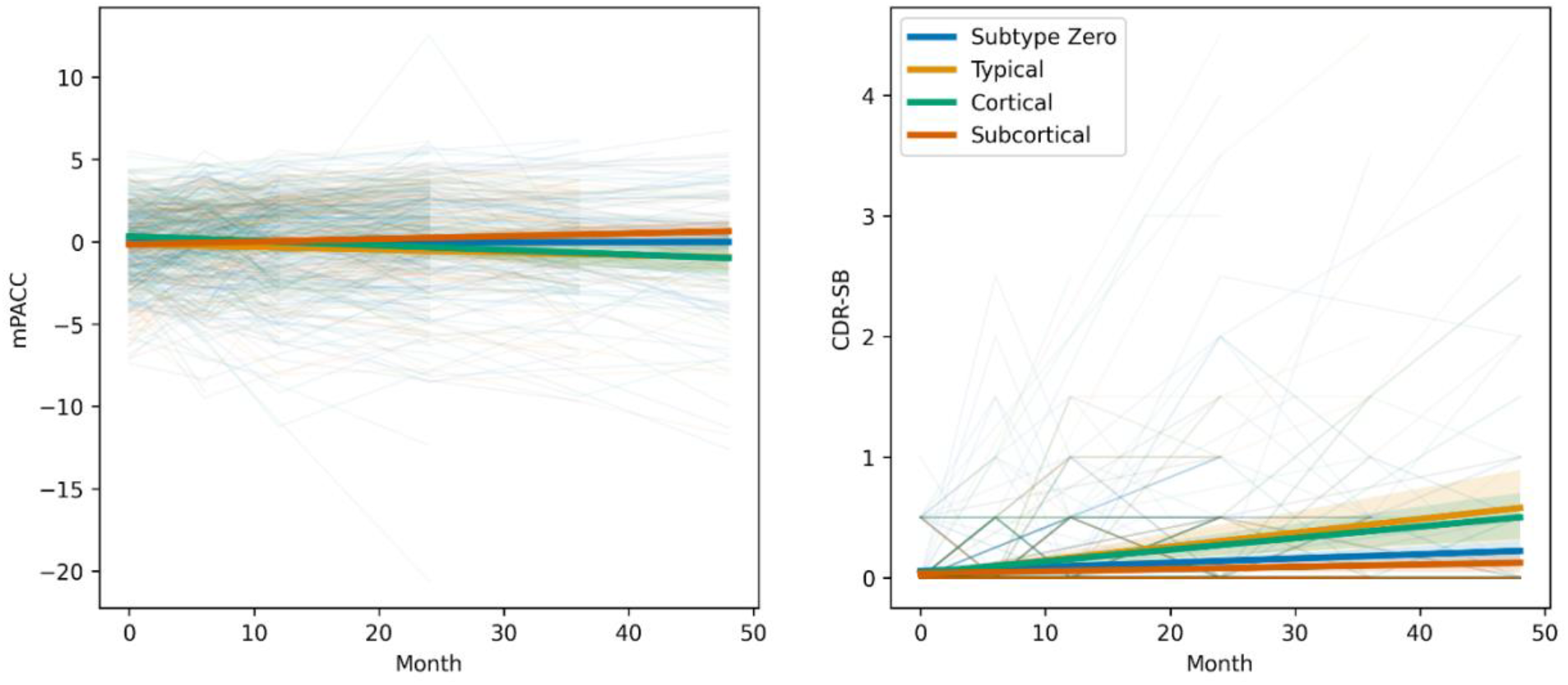
Analysis of A4 Study outcomes available in ADNI (CDR-SB and mPACC) over a period of four years, with individuals stratified by their baseline subtype assignment using the final disease progression model. Abbreviations: mPACC – Modified Preclinical Alzheimer Cognitive Composite; CDR-SB – Clinical Dementia Rating Sum of Boxes.

### Biological Heterogeneity Predicts Cognitive Heterogeneity

**Error! Reference source not found**. shows baseline PACC and CFI (total) scores in the A4 cohort stratified by subtype assignment. Broadly, baseline cognitive test scores were better in subtype zero and the *Subcortical* subtype. For the PACC score, there were significant differences between subtype zero (median = -0.013; IQR = -1.85 – 1.67) and both the *Typical* subtype (median = -0.78; IQR = -2.65 – 0.96; P = .004) and the *Cortical* subtype (median = -1.27; IQR = -3.34 – 0.83; P < .0001), but not for the *Subcortical* subtype (median = 0.03; IQR = -1.78 – 1.61; P = .87). Among the atrophy subtypes, PACC scores in both the *Typical* and *Cortical* subtypes were significantly different from those in the *Subcortical* subtype (P = .017 and P = .0006 respectively). For the CFI (total), there were also significant differences between subtype zero (median = 2.57; IQR = 1.04 – 5.00) and the *Typical* subtype (median = 3.02; IQR = 1.71 – 5.50; P = .04) and the *Cortical* subtype (median = 3.61; IQR = 1.67 – 6.65; P = .0003), but not the *Subcortical* subtype (median = 3.00, IQR = 1.14 – 5.58, P = .69). Among the atrophy subtypes, CFI did not differ statistically.

### Simulating A4 Study Subtype Cognitive Decline using ADNI

**Error! Reference source not found**. (lower panel) shows the distribution of subtype and staging for the ADNI subset selected using A4 inclusion criteria. A total of 731 individuals (726 CN, 5 MCI) from ADNI matched the criteria and had a baseline 3T scan. There were significant differences across in the subtypes in baseline age, sex, and amount of education, but not in APOE ε4 carriers or cognitive variables (eTable 1). Notably, each subtype had a much higher percentage of the female sex compared to subtype zero. The ADNI subset significantly differed from A4 in age and number of APOE ε4 alleles (but not sex or education) and had small but significant differences in the baseline MMSE and CDR-SB scores (eTable 2). Using the A4-trained SuStaIn model, these individuals were assigned a stage and subtype as follows: 390 (53.4%) were unclassifiable (subtype zero), 118 (16.1%) were *Typical*, 120 (16.4%) *Cortical*, and 103 (14.1%) *Subcortical*. The *Typical, Cortical*, and *Subcortical* subtypes had median stage assignments of 3 (IQR = 1 – 6), 3 (IQR = 2 – 5), and 4 (IQR = 2 – 7) respectively, similar to the A4 cohort.

**Error! Reference source not found**. shows cognitive decline measured by longitudinal scores over 4 years on mPACC (left) and CDR-SB (right) in the ADNI cohort, stratified by baseline subtype. Most subtypes showed minimal cognitive decline. For the mPACC, both the *Typical* (−0.23/yr; 95% CI, -0.41 to -0.05; P = .01) and *Cortical* (−0.24/yr; 95% CI, -0.42 to -0.06; P = .009) subtypes were associated with greater cognitive decline relative to subtype zero. For the CDR-SB, both the *Typical* (+0.09/yr; 95% CI, 0.06 to 0.12; P < .0001) and *Cortical* (+0.07/yr; 95% CI, 0.04 to 0.10; P < .0001) subtypes were associated with greater cognitive decline relative to subtype zero. Higher age at baseline was significantly associated with faster cognitive decline over the period for both CDR-SB (P = .04) and mPACC (P < .0001).

## Discussion

The A4 Study represents one of the largest studies of preclinical Alzheimer’s disease, providing an opportunity to leverage data-driven methods for analysis and potential applications, such as prediction of future decline and subgroup discovery. Using such a data-driven disease progression model on baseline MRI, the Subtype and Stage Inference (SuStaIn) algorithm uncovered previously undetected neurodegenerative heterogeneity in the A4 Study, despite enriching for amyloid (PET) positivity and extensive cognitive screening. Our DPM suggested three image-based subtypes in 523/1240 (42%) of participants, characterized as *Typical, Cortical*, and *Subcortical*, which share similarities with subtypes previously observed in cognitively impaired cohorts^5,16^. Our subtypes showed no significant differences in demographic, biomarker, or genetic variables which might otherwise be used to identify such differences.

The A4 cohort had 42% of individuals with sufficient neurodegeneration to be assigned to one of the three subtypes, which were approximately equal in size. Individuals within each subtype were spread across model stages (i.e. the extent of neurodegeneration), though the majority exhibited abnormality/atrophy in only a few regions, as expected by their preclinical status. This neurodegenerative heterogeneity was not captured by an amyloid and/or tau PET signal in the neocortex — we did not find significant differences between subtypes in the SUVr used for trial screening, and minimal differences across subtypes. Median amyloid and tau burden was greater in those assigned to a non-zero subtype, and numerically greatest in the *Cortical* subtype (not statistically significant for tau). This supports the notion that data-driven multivariate methods such as SuStaIn are uniquely useful for identifying (statistically significant) clinical heterogeneity in the absence of symptoms, above and beyond traditional screening tools.

Our image-based subtypes predicted cognitive heterogeneity, both cross-sectionally and longitudinally. Differences in baseline cognitive performance and longitudinal cognitive decline were both associated with DPM subtypes. In particular, the *Cortical* subtype displayed both poorer PACC scores at screening in A4 and more severe cognitive decline on mPACC and CDR-SB in ADNI (alongside the *Typical* subtype), suggesting that the screening protocol did not sufficiently capture (the variability in) underlying neurodegeneration. The ability to identify biological heterogeneity and predict cognitive heterogeneity supports the notion that image-based DPM can be used to inform clinical trial design, whether through screening (which could increase screen failures and initial costs), stratification, or covariate adjustment short of direct stratification^35^. Another potential design innovation could use image-based DPM to match subtypes to one of several targeted therapies in an umbrella design.

### Limitations

Our forecasts of cognitive decline were produced by combining MRI-based neurodegenerative heterogeneity in the A4 cohort with longitudinal data from an A4-like ADNI subset. These cohorts are not perfectly matched, despite using the A4 inclusion criteria to select participants from ADNI. We found small but statistically significant differences in the number of APOE ε4 alleles (more non-carriers in ADNI), age, baseline MMSE (0.34 points higher) and CDR-SB (0.02 points lower), indicating that the ADNI subset is at lower risk of AD decline than A4. This suggests that our longitudinal predictions may be underestimating effect sizes of decline (favoring better cognition), but only by testing directly on longitudinal data from A4 (when released) will our predictions be realized.

While longitudinal data is available in ADNI, only 160 (of 731) individuals had cognitive scores available after 4 years. This limits the sample size (particularly after stratification) and thus analysis, potentially reducing the translatability of our observations into the prospective outcomes of the A4 Study.

## Conclusions

The A4 Study represents a significant effort and investment into curating a preclinical cohort where anti-amyloid therapy may be more effective at slowing cognitive decline. Despite the study’s strict eligibility criteria and extensive screening, our disease progression model identified considerable neurodegenerative heterogeneity in 42% of participants, characterized by three MRI-based subtypes of neurodegeneration that can also be found in more advanced AD cohorts, associated with differences in cognition at baseline. Longitudinal analysis of an A4-matched subset of ADNI revealed significant differences in cognitive decline across the observed subtypes. These findings have potential ramifications for preclinical (and symptomatic) trials, where heterogeneity may obscure treatment effects. The use of data-driven disease progression modelling in trial design may increase statistical power of trials by accounting for the potential confounding effects of such heterogeneity on trial outcomes.

## Data Availability

All source data are available from the Image and Data Archive in the Laboratory Of NeuroImaging at the University of Southern California: https://ida.loni.usc.edu/. Derived data can be produced by running the processing pipelines described in the manuscript.

## Acknowledgements

NPO and CS acknowledge funding from a UKRI Future Leaders Fellowship (MR/S03546X/1). NPO, CS, FB and DCA acknowledge funding from the Early Detection of Alzheimer’s Disease Subtypes project (E-DADS; EU JPND 2019; MR/T046422/1). NPO, FB, and DCA acknowledge funding from the National Institute for Health Research University College London Hospitals Biomedical Research Centre. DCA also acknowledges support from Wellcome Trust award 221915/Z/20/Z. PJM is supported by EU-EFPIA Innovative Medicines Initiative 2 Joint Undertaking (IMI 2 JU) Amyloid Imaging to Prevent Alzheimer’s Disease (AMYPAD, grant agreement number: 115952). DMC is supported by the UK Dementia Research Institute which receives its funding from DRI Ltd, funded by the UK Medical Research Council, Alzheimer’s Society and Alzheimer’s Research UK (ARUK-PG2017-1946), the UCL/UCLH NIHR Biomedical Research Centre, and the UKRI Innovation Scholars: Data Science Training in Health and Bioscience (MR/V03863X/1). MCD acknowledges funding from Eli Lilly and Company, Alzheimer’s Association, and NIH (R01AG063689 and U24AG057437).

Data used in preparation of this article were obtained from the Alzheimer’s Disease Neuroimaging Initiative (ADNI) database (adni.loni.usc.edu). As such, the investigators within the ADNI contributed to the design and implementation of ADNI and/or provided data but did not participate in analysis or writing of this report. A complete listing of ADNI investigators can be found at: http://adni.loni.usc.edu/wp-content/uploads/how_to_apply/ADNI_Acknowledgement_List.pdf. Data collection and sharing for this project was funded by the Alzheimer’s Disease Neuroimaging Initiative (ADNI) (National Institutes of Health Grant U01 AG024904) and DOD ADNI (Department of Defense award number W81XWH-12-2-0012).

The A4 Study is a secondary prevention trial in preclinical Alzheimer’s disease, aiming to slow cognitive decline associated with brain amyloid accumulation in clinically normal older individuals. The A4 Study is funded by a public-private-philanthropic partnership, including funding from the National Institutes of Health-National Institute on Aging, Eli Lilly and Company, Alzheimer’s Association, Accelerating Medicines Partnership, GHR Foundation, an anonymous foundation and additional private donors, with in-kind support from Avid and Cogstate. The companion observational Longitudinal Evaluation of Amyloid Risk and Neurodegeneration (LEARN) Study is funded by the Alzheimer’s Association and GHR Foundation. The A4 and LEARN Studies are led by Dr. Reisa Sperling at Brigham and Women’s Hospital, Harvard Medical School and Dr. Paul Aisen at the Alzheimer’s Therapeutic Research Institute (ATRI), University of Southern California. The A4 and LEARN Studies are coordinated by ATRI at the University of Southern California, and the data are made available through the Laboratory for Neuro Imaging at the University of Southern California. The participants screening for the A4 Study provided permission to share their de-identified data in order to advance the quest to find a successful treatment for Alzheimer’s disease. We would like to acknowledge the dedication of all the participants, the site personnel, and all of the partnership team members who continue to make the A4 and LEARN Studies possible. The complete A4 Study Team list is available on: a4study.org/a4-study-team.

## Supplementary Material

### eMethods 1: Alzheimer’s Disease Neuroimaging Initiative (ADNI)

Data used in the preparation of this article were obtained from the Alzheimer’s Disease Neuroimaging Initiative (ADNI) database (adni.loni.usc.edu). The ADNI was launched in 2003 as a public-private partnership, led by Principal Investigator Michael W. Weiner, MD. The primary goal of ADNI has been to test whether serial magnetic resonance imaging (MRI), positron emission tomography (PET), other biological markers, and clinical and neuropsychological assessment can be combined to measure the progression of mild cognitive impairment (MCI) and early Alzheimer’s disease (AD). For up-to-date information, see www.adni-info.org. MRI scans were downloaded from LONI on 27^th^ February 2022.

### eMethods 2: A4 Amyloid & Tau PET Protocols

Florbetapir PET scans were collected from 50–70 minutes post-injection and were generally reconstructed in 4×5-minute frames, which were aligned and averaged into a single 3D NIFTI-formatted image suitable for SUVr analysis. Flortaucipir PET scans were collected 90-110 minutes post-injection. The preprocessing/alignment was performed by the A4 Study investigators before data release, and so the AmyPET pipeline implemented here did not include the preprocessing of raw PET count data.

**eTable 1.**
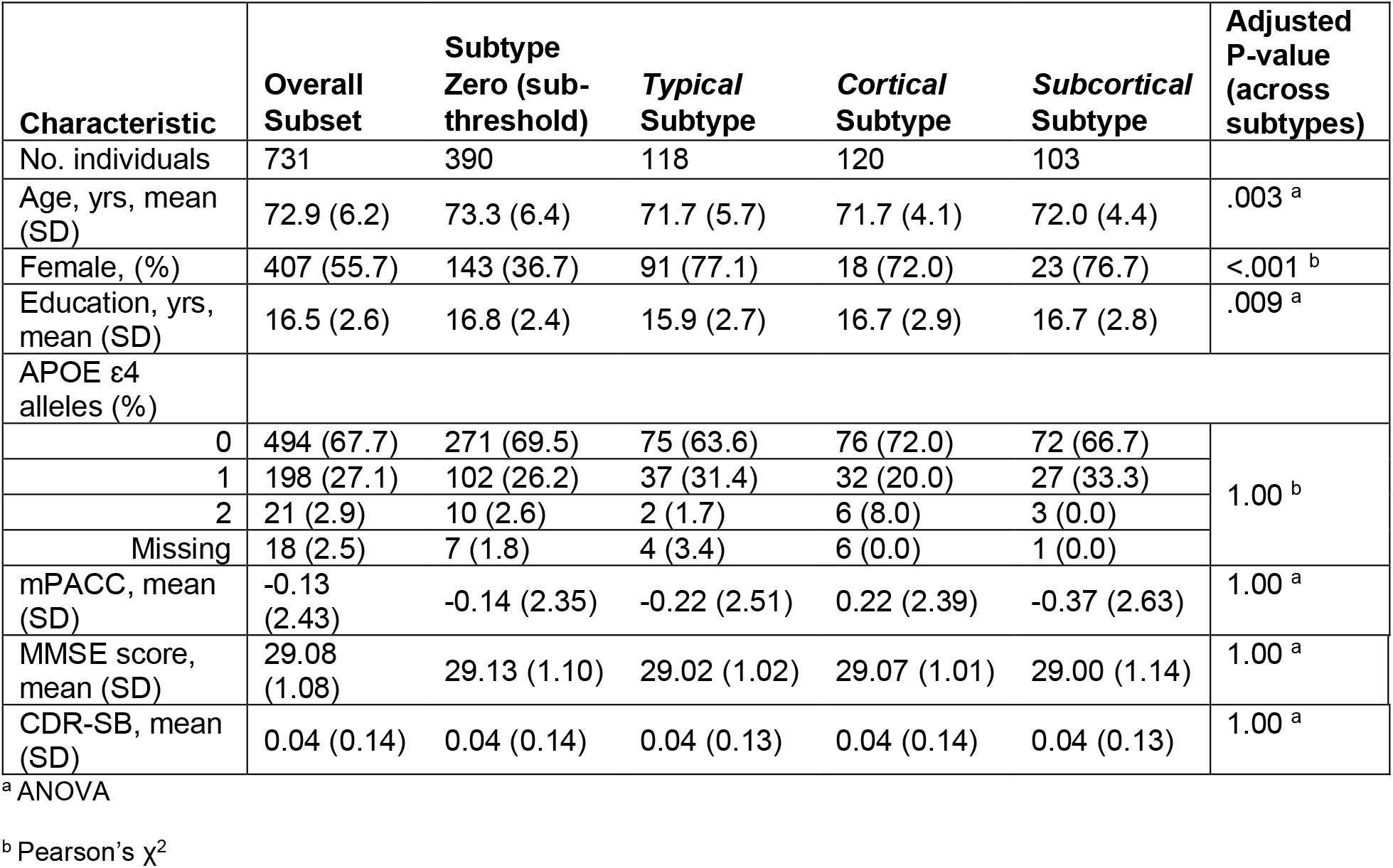
Characteristics of the ADNI subset selected using the A4 Study inclusion criteria. The subset was assigned a subtype by the A4-trained 3-subtype model. Differences across the subtypes were assessed using either an ANOVA or Pearson’s χ^2^ test (following Holm-Bonferroni adjustment for multiple comparisons). There was a significant difference between sex across the subtypes, but no other variables. Abbreviations: mPACC – Modified Preclinical Alzheimer Cognitive Composite; MMSE – Mini-Mental State Examination; CDR-SB – Clinical Dementia Rating Sum of Boxes

**eTable 2.**
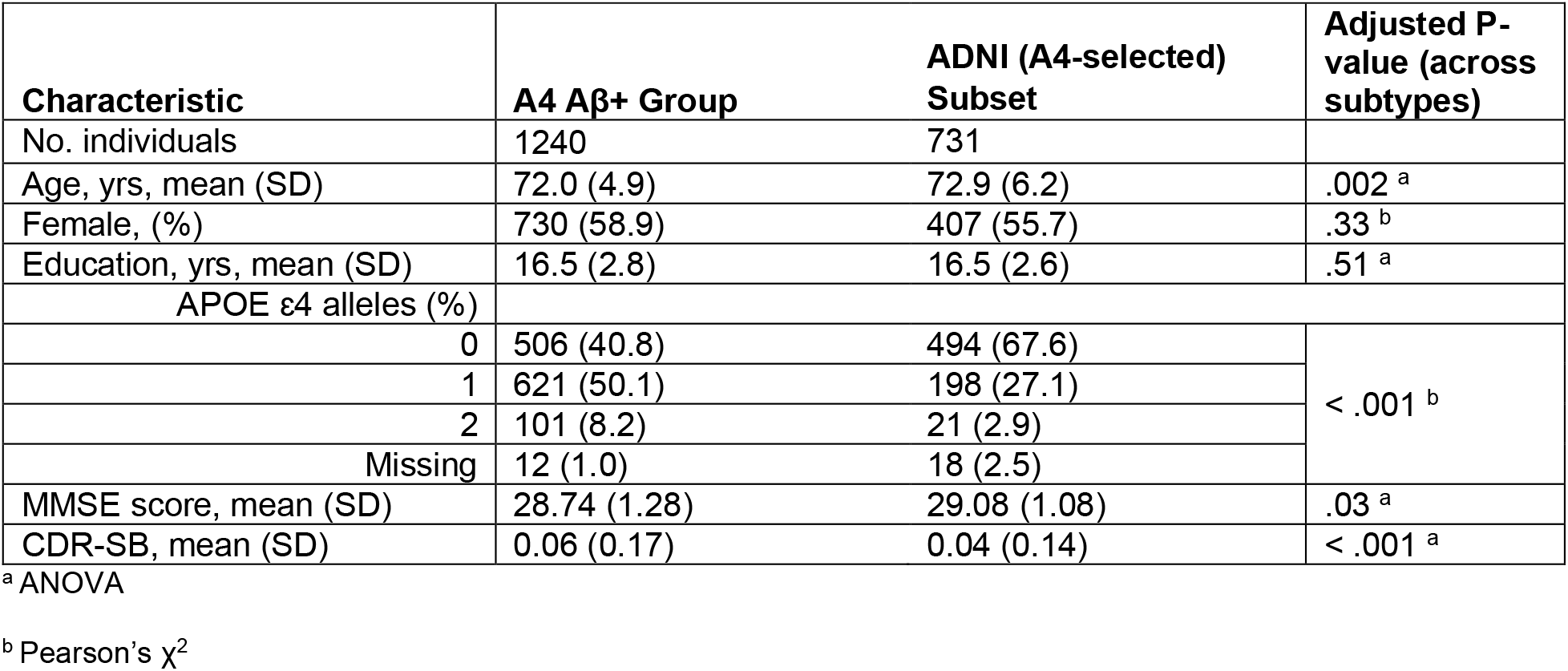
Comparison of A4 Aβ+ group and the ADNI subset selected using the A4 Study inclusion criteria. Differences between the cohorts were assessed using either an ANOVA or Pearson’s χ^2^ test (following Holm-Bonferroni adjustment for multiple comparisons) on variables available in both cohorts relevant to this study. Abbreviations: MMSE – Mini-Mental State Examination; CDR-SB – Clinical Dementia Rating Sum of Boxes

